# Performance and Operational Evaluation of the Access Bio CareStart Rapid Antigen Test in a High-throughput Drive-through Community Testing Site in Massachusetts

**DOI:** 10.1101/2021.03.07.21253101

**Authors:** Nira R. Pollock, Kristine Tran, Jesica R. Jacobs, Amber E. Cranston, Sita Smith, Claire Y. O’Kane, Tyler J. Roady, Anne Moran, Alison Scarry, Melissa Carroll, Leila Volinsky, Gloria Perez, Pinal Patel, Stacey Gabriel, Niall J. Lennon, Lawrence C. Madoff, Catherine Brown, Sandra C. Smole

## Abstract

**Background:** To facilitate deployment of point-of-care testing for SARS-CoV-2, we evaluated the Access Bio CareStart COVID-19 Antigen test in a high-throughput, drive-through, free community testing site using anterior nasal (AN) swab RT-PCR for clinical testing.

**Methods:** Consenting symptomatic and asymptomatic children (≤18 years) and adults received dual AN swabs. CareStart testing was performed with temperature/humidity monitoring. All tests had two independent reads to assess inter-operator agreement. Patients with positive CareStart results were called and instructed to isolate pending RT-PCR results. The paired RT-PCR result was the reference for sensitivity and specificity calculations.

**Results:** Of 1603 participants, 1245 adults and 253 children had paired RT-PCR/CareStart results and complete symptom data. 83% of adults and 87% of children were asymptomatic. CareStart sensitivity/specificity were 84.8% (95% confidence interval [CI] 71.1-93.7)/97.2% (92.0-99.4) and 85.7% (42.1-99.6)/89.5% (66.9-98.7) in adults and children, respectively, within 5 days of symptoms. Sensitivity/specificity were 50.0% (41.0-59.0)/99.1% (98.3-99.6) in asymptomatic adults and 51.4% (34.4-68.1)/97.8% (94.5-99.4) in asymptomatic children. Sensitivity in all 234 RT-PCR-positive people was 96.3% with cycle threshold (Ct) ≤25, 79.6% with Ct ≤30, and 61.4% with Ct ≤35. All 21 false positive CareStart tests had faint but normal bands. Inter-operator agreement was 99.5%. Operational challenges included identification of faint test bands and inconsistent swab elution volumes.

**Conclusions:** CareStart had high sensitivity in people with Ct ≤25 and moderate sensitivity in symptomatic people overall. Specificity was unexpectedly lower in symptomatic versus asymptomatic people. Excellent inter-operator agreement was observed, but operational challenges indicate that operator training is warranted.

## Introduction

Although nucleic acid amplification tests (NAATs) for SARS-CoV-2 can be highly sensitive and are being performed at high volumes in centralized laboratories worldwide [1, 2], global testing capacity and turnaround time remain insufficient. The benefits of decentralized and expedited testing have driven the development of rapid diagnostic tests (RDTs) for point-of-care (POC) use that detect SARS-CoV-2 nucleocapsid antigen (Ag) in as little as 10-15 minutes. As of February 10, 2020, there are 11 Ag RDTs with FDA Emergency Use Authorization (EUA) [3] that can be performed by personnel without formal laboratory training in patient care settings that operate under a Clinical Laboratory Improvement Amendments (CLIA) Certificate of Waiver.

Several tests that are visually read have demonstrated consistently high specificity (>99%) in field testing and high sensitivity in individuals with high viral burden (typically defined by the surrogate measure of a low cycle threshold [Ct] value in a real-time reverse transcription polymerase chain reaction [RT-PCR] assay performed on a separate swab collected in parallel) [4–10]. However, variability in sensitivity estimates yielded from field studies of individual Ag RDTs (e.g., the Abbott BinaxNOW COVID-19 Ag Card, [6, 8, 10]) have reinforced the fact that the performance of an Ag RDT must be established in the settings, conditions, and populations of intended use. Although nasopharyngeal (NP) sampling remains the reference method, anterior nasal (AN) sampling substantially increases testing access and acceptability, and a recent comparison study showed that sensitivity with self-collected nasal mid-turbinate swabs versus professionally-collected NP swab samples was similar [11].

The Access Bio CareStart COVID-19 Antigen (Ag) test has FDA EUA for AN swab samples [12] and can provide visually-read results at POC in 10 minutes. The potential for use of this test at large scale, and the paucity of data for test performance in asymptomatic individuals and in children, motivated us to perform an implementation and performance evaluation in a high-volume, high-prevalence community testing site currently using AN swab RT-PCR for clinical testing.

## Methods

### Study population

The study was performed from January 11-January 22, 2021, at the Lawrence General Hospital “Stop the Spread” drive-through testing site, which accommodates Massachusetts residents from the surrounding area. CareStart testing was performed under the site’s CLIA waiver. No study-specific effort was made to recruit individuals to present to the testing site. Two of seven drive-through lanes were utilized for the study. Verbal consent for dual AN swabbing was obtained from adults and guardians of minors (with verbal assent for ages 7-17). Participants were informed of the Ag RDT results reporting plan (below). Presence or absence of symptoms (sore throat, cough, chills, body aches, shortness of breath, fever, runny nose, congestion, nausea, vomiting, diarrhea, loss of taste or smell) was recorded for each participant, including the date of symptom onset. Participants whose symptoms started on the day of testing were classified as Day 0. The study was reviewed by the Massachusetts Department of Public Health IRB and deemed not human subjects research.

### Swab collection procedure

Cars with consented patients were marked with a glass marker, notifying the specimen collector to collect two AN swabs rather than one. Swab collection details are in Supplementary Methods; in brief, collection involved swabbing both nostrils with each swab and operators alternated which swab was collected first (for RT-PCR vs. CareStart). CareStart swabs were captured in an empty sterile tube and taken to the testing trailer by a designated “runner.” Time of sample collection was recorded, and CareStart tests were initiated within an hour of collection.

### Access Bio CareStart COVID-19 Antigen test performance

The test was performed by trained operators (Master’s or PhD level laboratorians) according to the manufacturer instructions for use (IFU) [12]; note that testing of individuals with symptoms >5 days or without symptoms is off-label. Details of kit storage, quality control, and testing and results reporting procedures are in Supplementary Methods.

### RT-PCR assay

Dry AN swabs were collected per site routine and transported at room temperature to the Broad Institute for testing using the CRSP SARS-CoV-2 Real-time Reverse Transcriptase (RT)-PCR Diagnostic Assay under EUA [13]. Details are in Supplementary Methods.

### Results reporting

All positive CareStart results were verbally reported by phone to individuals the same day by the Massachusetts Department of Public Health (DPH). Participants with positive CareStart results were informed that the Ag result was presumptive positive and that they should isolate until they received their confirmatory RT-PCR result in 1-2 days. If the RT-PCR result was negative, they could discontinue isolation. RT-PCR results were provided to the patient by the Lawrence General Hospital’s portal or by walk up to a designated location at the Lawrence General Hospital. RT-PCR results were reported to DPH through routine electronic laboratory reporting mechanisms and individuals with positive results were referred to local boards of health or the Community Tracing Collaborative for instruction on isolation and case investigation for contact tracing.

### Statistical analysis

Sensitivity, specificity, negative predictive value (NPV), and positive predictive value (PPV) for the CareStart test were calculated using the RT-PCR result as the reference. 95% CI were calculated using the Clopper-Pearson method. Analyses utilized Microsoft Excel and GraphPad Prism.

## Results

Two different lots of CareStart kits were used for the study. Each operator was able to set up and read 20 tests per hour, and two operators were able to manage testing of samples coming from two drive-through lanes. 1493/1498 (99.7%) tests were initiated within 1 hour of collection (a window approved by the test manufacturer prior to study start); the median interval between sample collection and test initiation was 31 minutes (range 12–103 min) and the 5 tests performed at ≥1 hour were all negative (both Ag and paired RT-PCR results). All tests were read within the requisite 5 minute window per the EUA IFU [12]. Temperature and humidity in the testing trailer (Supplementary Methods) from 7:30AM-6:00PM ranged from 70.5-74.3°F and 11.7-40.9%, respectively. All testing and kit storage temperatures met manufacturer recommendations [12].

Of 1603 participants [excluding those with invalid or missing RT-PCR results (n=48) and those with missing clinical data (n=57)], 1498 had paired RT-PCR/CareStart results and complete symptom data, including 221 asymptomatic children, 1036 asymptomatic adults, 32 symptomatic children, and 209 symptomatic adults. Symptomatic individuals were further classified by days (D) since symptom onset; both cutoffs of ≤5D and ≤7D of symptoms were evaluated given that the CareStart test EUA is for individuals within 5D of symptom onset [12], but 7D is a window used by several other commercial Ag RDTs [3]. Clinical data for the study population are presented in Table 1 (demographics) and Supplementary Tables 1 and 2 (symptoms).

**Table 1.** Clinical characteristics of adult and pediatric patients contributing paired samples.

|  | **Adults,**<br>**symptomatic**<br>**(n=209)** | **Adults,**<br>**asymptomatic**<br>**(n=1036)** | **Children,**<br>**symptomatic**<br>**(n=32)** | **Children,**<br>**asymptomatic**<br>**(n=221)** |
| --- | --- | --- | --- | --- |
| **Age in years, n**<br>**(%)** |  |  |  |  |
| <7 | n/a | n/a | 13 (40.6) | 60 (27.2) |
| 7-13 | n/a | n/a | 12 (37.5) | 73 (33.0) |
| 14-18 | n/a | n/a | 7 (21.9) | 88 (39.8) |
| 19-29 | 58 (27.8) | 313 (30.2) | n/a | n/a |
| 30-49 | 102 (48.8) | 381 (36.8) | n/a | n/a |
| 50-69 | 42 (20.1) | 290 (28.0) | n/a | n/a |
| 70 and older | 7 (3.3) | 52 (5.0) | n/a | n/a |
| **Sex, % female** | 57.4 | 53.0 | 56.3 | 53.4 |
| **Days of**<br>**symptoms prior**<br>**to COVID-19**<br>**test, median**<br>**(IQR)** | 3 (2,6)<sup>a</sup> | n/a | 3 (2,4)<sup>b</sup> | n/a |

IQR, interquartile range; n/a, not applicable
<sup>a</sup>Range 0-44 days
<sup>b</sup>Range 1-20 days

### CareStart performance in adults and children (≤18 years old)

Sensitivity, specificity, PPV and NPV calculations for CareStart results vs. RT-PCR results as the reference, for each clinical subgroup, are presented in Table 2. Tables with data for each subgroup are presented in Supplementary Table 3.

**Table 2.** Performance of the Access Bio CareStart versus RT-PCR (reference method) for detection of SARS-CoV-2 in anterior nasal swab samples from adults and children (≤18)

| Age | Group | N | Prevalence | Sensitivity<br>(95% CI) | Specificity<br>(95% CI) | PPV<br>(95% CI) | NPV<br>(95% CI) |
| --- | --- | --- | --- | --- | --- | --- | --- |
| All | Sx + ASx | 1498 | 15.6% | 57.7%<br>(51.1 – 64.1) | 98.3%<br>(97.5 – 99.0) | 86.5%<br>(80.6 – 90.9) | 92.6%<br>(91.5 – 93.6) |
| Pediatric | Sx + ASx | 253 | 18.2% | 56.5%<br>(41.1 – 71.1) | 96.6%<br>(93.2 – 98.6) | 78.8%<br>(63.2 – 88.9) | 90.9%<br>(87.8 – 93.3) |
| Adult | Sx + ASx | 1245 | 15.1% | 58.0%<br>(50.6 – 65.1) | 98.7%<br>(97.8 – 99.3) | 88.6%<br>(82.0 – 93.0) | 93.0%<br>(91.8 – 94.0) |
| Pediatric | ASx | 221 | 16.7% | 51.4%<br>(34.4 – 68.1) | 97.8%<br>(94.5 – 99.4) | 82.6%<br>(63.2 – 92.9) | 90.9%<br>(87.8 – 93.3) |
| Adult | ASx | 1036 | 12.4% | 50.0%<br>(41.0 – 59.0) | 99.1%<br>(98.3 – 99.6) | 88.9%<br>(79.8 – 94.2) | 93.3%<br>(92.2 – 94.3) |
| Pediatric | Sx ≤5D | 26 | 26.9% | 85.7%<br>(42.1 – 99.6) | 89.5%<br>(66.9 – 98.7) | 75.0%<br>(43.8 – 92.0) | 94.5%<br>(73.4 – 99.1) |
| Adult | Sx ≤5D | 152 | 30.3% | 84.8%<br>(71.1 – 93.7) | 97.2%<br>(92.0 – 99.4) | 92.9%<br>(80.9 – 97.6) | 93.6%<br>(88.1 – 96.7) |
| Pediatric | Sx ≤7D | 27 | 25.9% | 85.7%<br>(42.1 – 99.6) | 85.0%<br>(62.1 – 96.8) | 66.6%<br>(40.3 – 85.6) | 94.5%<br>(73.4 – 99.1) |
| Adult | Sx ≤7D | 169 | 30.2% | 84.3%<br>(71.4 – 93.0) | 97.5%<br>(92.8 – 99.5) | 93.5%<br>(82.4 – 97.8) | 93.5%<br>(88.4 – 96.5) |
| Pediatric | Sx >7D | 5 | 40.0% | 50.0%<br>(1.3 – 98.7) | 100.0%<br>(29.2 – 100.0) | 100.0% | 75.0%<br>(42.9 – 92.3) |
| Adult | Sx >7D | 40 | 22.5% | 22.2%<br>(2.8 – 60.0) | 90.3%<br>(74.3 – 98.0) | 40.0%<br>(11.6 – 77.3) | 80.0%<br>(73.5 – 85.3) |

CI, confidence interval; PPV, positive predictive value; NPV, negative predictive value; Sx, symptomatic; ASx, asymptomatic; D, days

Sensitivity in adults with symptoms ≤5D was 84.8%, similar to that in the CareStart IFU (87.2%) [12]. Sensitivity in children with symptoms ≤5D was 85.7%. Specificity in these symptomatic adults and children were 97.2% and 89.5%, respectively. Relative to symptomatic individuals, sensitivity in asymptomatic adults and children was lower at 50.0% and 51.4%, respectively, while specificity was higher (99.1% and 97.8%, respectively).

### Discordant analysis and analysis of Ct values

There were 21 false positive CareStart results across all 1498 individuals tested, including 8 asymptomatic adults, 4 asymptomatic children, 3 children with symptoms ≤7D, 3 adults with symptoms ≤7D, and 3 adults with symptoms >7D. All 21 false positives were scored as faint bands by both independent readers, and there was nothing unusual noted about band morphology. One in 21 swabs and two in 21 swabs had minimal and moderate blood, respectively; no excess mucus was observed. Ninety-nine false negative CareStart results were found in 1498 individuals; 5/99 swabs had been minimally bloody and 1/99 had excess mucous. No correlation was observed between false positive or false negative CareStart results and presence of symptoms of congestion/rhinorrhea, order of swabbing (Ag vs. RT-PCR), time between sample collection and test initiation, or test kit lot.

Distributions of Ct values for RT-PCR positive symptomatic (by days post symptom onset) and asymptomatic children and adults are shown in Figure 1; false negative vs. true positive paired CareStart results are indicated for all individuals. As expected, false negative CareStart results were paired with RT-PCR tests with higher Ct values. Sensitivity was evaluated at three different Ct cutoffs: ≤25, ≤30, ≤35 (Supplementary Table 4). Sensitivity in all subgroups combined (n=234 RT-PCR-positive individuals) was 96.3% with Ct ≤25, 79.6% with ≤30, and 61.4% with ≤35. Band strength (1 = faint, n = 45; 2 = medium, n = 11; 3 = strong, n = 79) as interpreted by the primary reader for the 135 true positive CareStart tests correlated clearly with Ct value, with median Ct (IQR) of 24.7 (21.8-27.8), 22.8 (19.9-24.5), and 18.0 (15.7-20.4), respectively, as shown in Figure 2. Five of 99 false negative CareStart results had a paired RT-PCR Ct ≤25 (1 child, 4 adults), and 43/99 had paired Ct ≤30; the median (IQR) Ct value for the 99 individuals with false negative CareStart results was 32.0 (28.7-34.5). Distribution of the 99 false negative results among clinical subgroups is shown in the 2×2 tables in Supplementary Table 3.

**Figure 1.**
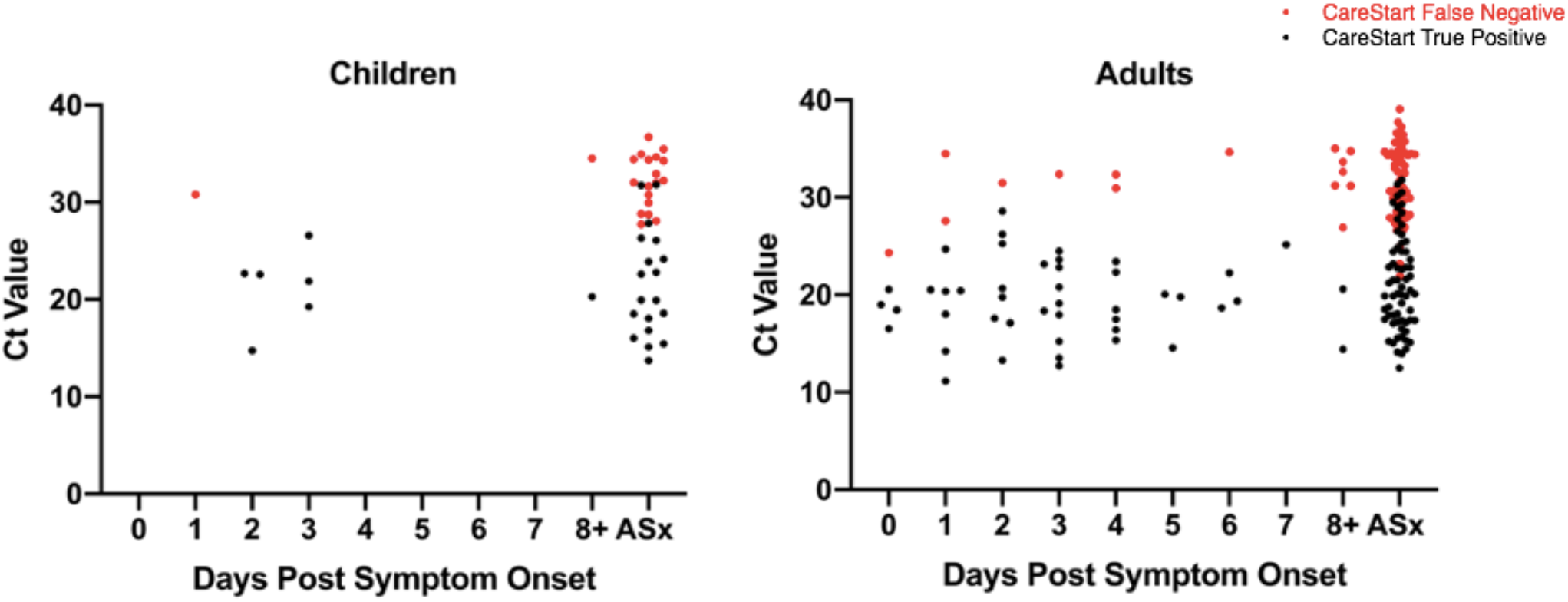
Distribution of Cycle Threshold (Ct) Values in RT-PCR-positive Children and Adults by Days Post Symptom Onset. Ct values for each RT-PCR-positive individual are shown; red circles indicate false negative Access Bio CareStart results and black circles, true positive CareStart results. Participants whose symptoms started on the day of testing are indicated as Day 0. RT-PCR, real-time reverse transcription polymerase chain reaction; ASx, asymptomatic.

**Figure 2.**
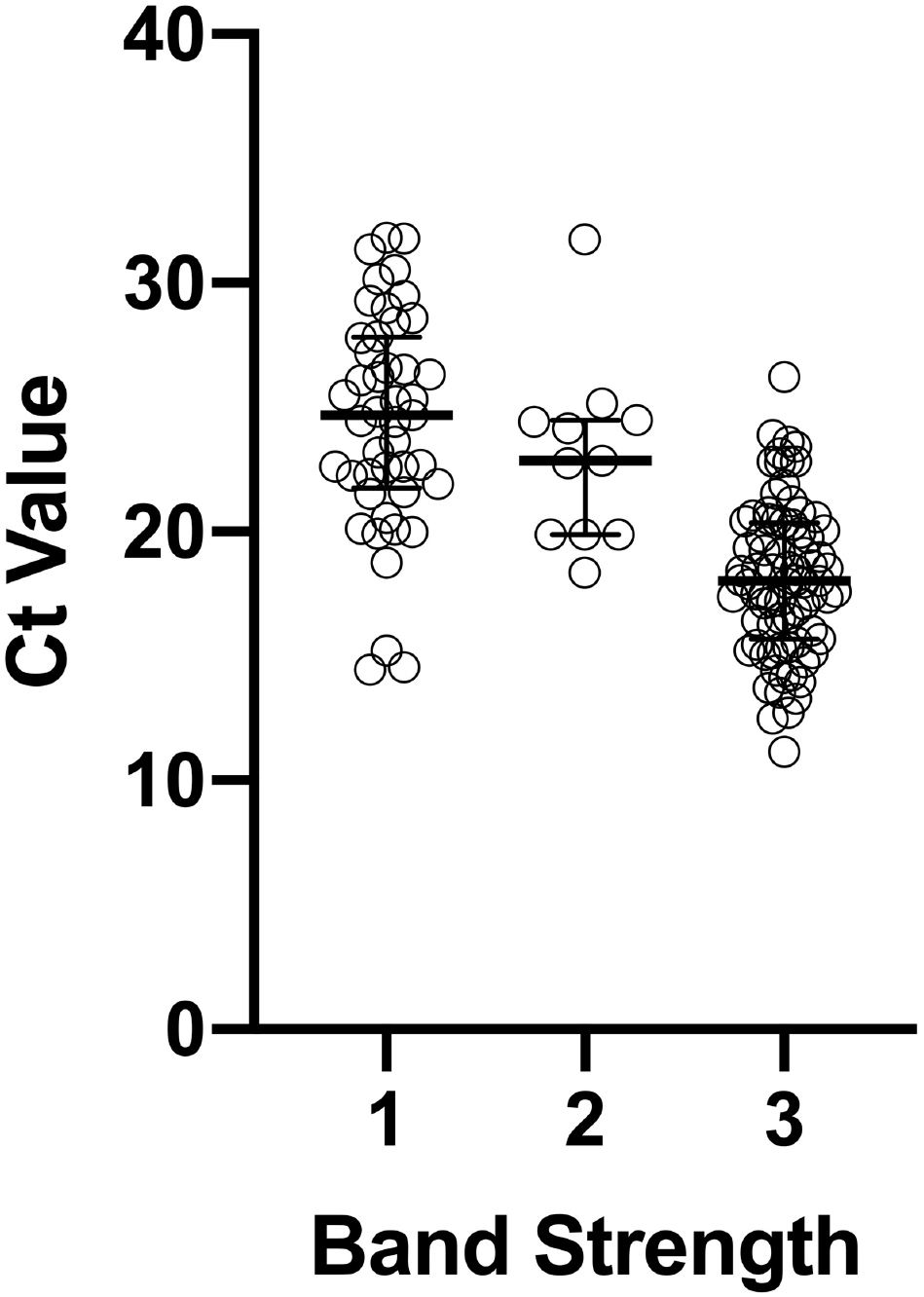
Correlation of Cycle Threshold (Ct) value with CareStart band strength. Correlation of Access Bio CareStart test band strength (1= faint, n = 45; 2 = medium, n = 11; 3 = strong, n = 79, as interpreted by the first reader) and RT-PCR Cycle Threshold (Ct) value from a swab collected in parallel are shown for the 135 individuals with true positive CareStart tests. Median Ct values (IQR) were 24.7 (21.8-27.8), 22.8 (19.9-24.5), and 18.0 (15.7-20.4) for Groups 1, 2, and 3, respectively.

### Operational findings

Inter-operator agreement was excellent, with the two readers agreeing on the positive versus negative result for 1490/1498 (99.5%) CareStart tests. The 8 discordant reads were all faint positive vs. negative. The two readers disagreed on the strength of the positive band (faint vs. medium vs. strong) in 7 cases. Overall, readers noted that detection of faint positive bands and distinguishing them from negative results required very close observation and good lighting, particularly due to the blue color of a faint positive band potentially resembling a shadow. A subset of 10 extracted samples with positive CareStart results were retested four hours later (as per the IFU [12], samples can sit for up to 4 hours in extraction buffer before testing) and all 10 repeat results were qualitatively the same as the original results.

Operators noted that the volume of extraction buffer absorbed by the swab head was inconsistent, and that it was sometimes difficult to elute sufficient volume from the head of the swab for testing (a process that requires squeezing the sides of the extraction vial [12]). This issue required careful observation and, ultimately, experience to overcome. Additionally, operators noted that the polyester swab head did not seem completely stable (occasional apparent unravelling of the head surface, at an anecdotal rate of up to 5/200 tests per day). Coincidentally, the same swab brand was already in use for RT-PCR testing at this site, making it possible to confirm that this “unravelling” at the time of patient swabbing had already been observed over an extended time period with this particular swab. The operators found that a good deal of force was required to fit the caps on the extraction vials, and that the cap did not “click” as per the IFU, leading to concerns about spillage; they also found that peeling the foil off of the extraction vial was difficult and sometimes led to dripping of buffer and slippery gloves. Each skilled laboratorian was able to perform and read ∼20 tests per hour; operators felt that throughput was limited by the short read time window (5’). No invalid CareStart test results were observed.

## Discussion

The development of Ag RDTs offers the opportunity to dramatically expand COVID-19 testing capacity and also raises critical questions about how these tests could and should be used. Field evaluation of an Ag RDT at POC in the settings and populations of intended use can add tremendously to the performance data available in manufacturer package inserts and guide test deployment. Gaps in performance data, particularly test performance in asymptomatic adults and both symptomatic and asymptomatic children, must be filled in order to optimally deploy Ag RDTs.

Prior to this study, only minimal data for performance of the CareStart test in symptomatic individuals was available in the manufacturer’s IFU [12]. In order to understand how well the CareStart RDT could perform in both symptomatic and asymptomatic adults and children in a real-world but also best-case testing scenario, we implemented the test at a high-volume community testing site already experienced in collecting AN samples for RT-PCR. The CareStart test was performed by trained laboratory personnel, with careful attention paid to sample collection, results documentation, and quality control.

We found that the CareStart test had high sensitivity in individuals with highest viral burden (96.3% sensitive with paired PCR Ct value ≤25) and moderate sensitivity (84.8/85.7%) in symptomatic adults/children (≤5D of symptoms), respectively (acceptable per the FDA’s target of ≥80% [14, 15]). Sensitivity in symptomatic individuals with ≤5D of symptoms (the time frame recommended in the EUA IFU; [12]) was comparable to sensitivity in those with ≤7D of symptoms (the time frame recommended for some other Ag RDTs like BinaxNOW[16]). Sensitivity in asymptomatic adults and children was substantially lower than that in symptomatic individuals, which may correspond with the broad viral load distribution observed in this population (likely capturing early and late infections given unknown disease onset). Thus, the test does not appear to be optimal for ruling out SARS-CoV-2 infection in asymptomatic adults or children; use in serial testing programs and for testing of contacts of known cases deserves independent study. FDA does provide guidance for consideration of serial Ag testing if the sensitivity is lower, e.g., 70% [14, 15].

It should be noted that in all groups, CareStart sensitivity followed Ct value distribution, with 96.3% sensitivity observed in all participants with Ct ≤25 and 79.6% in those with Ct ≤30. Although false negative CareStart results were largely confined to those perhaps least likely to transmit SARS-CoV-2, the sensitivity of the CareStart test by Ct threshold cutoff was lower than observed in our recent study of the Abbott BinaxNOW in the same testing site [6] (99.3% with Ct ≤25, 95.8% with ≤30, and 81.2% with ≤35). For the RT-PCR assay used in this study, Ct values of 25, 30, and 35 correspond to approximately 5.4×10^5^, 1.7×10^4^, and 5.5×10^2^ copies/mL, respectively (Niall Lennon, personal communication; Supplementary Methods). The sources of the lower CareStart sensitivity are unknown, but one clear possibility is the dilution of the swab in extraction buffer in the CareStart test format.

Unexpectedly, we found that specificity of the CareStart test was lower in symptomatic people than in asymptomatic people: specificity in adults/children within 5D of symptoms were 97.2%/89.5% and in asymptomatic adults/children were 99.1%/97.8%, respectively. This pattern was not observed in our BinaxNOW study (100% specificity in people within 7D of symptoms, and 99.6%/99.0% specificity in asymptomatic adults/children, respectively [6]) nor in the Access Bio prospective AN swab study detailed in the CareStart EUA IFU (100% specificity in symptomatic individuals [12]). This specificity is also lower than that observed in a number of other field studies of visually-read Ag RDTs, including the BinaxNOW, SD Biosensor SD Q, and Abbott PanBio RDTs (>99% for all, [4–10]). This finding might suggest a pre-analytical issue unique to this test or to this study (e.g., mucus on the swab, or the swab itself), but we did not see any obvious overrepresentation of either nasal congestion/rhinorrhea or visible blood/mucus on the swabs of those with false positive results. Cross-reactivity with another pathogen in symptomatic patients is another possible explanation. No unusual band morphologies were noted in the 21 false positive results, and all were faint positive bands. We noted occasional “unravelling” of the swab head which might have contributed; because this issue was infrequent, we did not document when it occurred and thus are unable to correlate this event with false positive results. The overall variability we observed in absorption of extraction buffer by the swab head and subsequent elution volume may or may not have contributed to lower specificity. We note that the swab used for this study [SteriPack Sterile Polyester Spun Swab, 3” (Lakeland, Florida)] is the same swab that was used in the CareStart EUA study and will be included in the AN kit going forward. This same swab has been consistently used for RT-PCR testing over the past year at this site, with the same occasional “unravelling” noted at the time of sample collection, indicating that this does not appear to be a lot issue. Logistics of sample collection and testing in high volume at the site did require a window of time between collection and testing (median 31 minutes), but it was not possible to put each swab “immediately” into extraction buffer as stated in the IFU [12], and our window of one hour between collection and testing was pre-approved by the test manufacturer. Test specificity will need further confirmation in future studies.

Our study yielded some important operational findings relevant to test implementation. Inter-operator agreement on positive/negative results was near 100%, confirming that only one person is needed to read each test result. The main challenge to reading the test was distinguishing a faint positive band from a negative result; operators attributed this in part to the blue color of the faint positive band resembling a shadow, and recommended use of a strong light source in close proximity to the test device during test reading.

The requirement for extraction of the swab in buffer introduced multiple operational challenges. The volume of extraction buffer absorbed by the swab head appeared to be inconsistent, and operators sometimes had difficulty eluting sufficient volume from the head of the swab for testing [by squeezing the sides of the extraction vial as per the IFU; [12]]. This issue required careful observation and over time became easier for the operators. The occasional unravelling of the swab head in buffer (anecdotally, up to 5/200 tests/day) is described above. Operators noted that it was difficult to peel the foil off the extraction vial while wearing gloves. Additionally, a residual, small drop of buffer on the inner lid of the foil sometimes made gloves slippery with buffer, and operators had difficulty fitting the caps securely on the extraction vials, both of which led to concern about dropping vials during the extraction step. Each skilled laboratorian in the study was able to perform and read ∼20 tests per hour; although the test only takes 10’ to perform, the short read time window (5’) required frequent breaks in test setup and thus decreased throughput.

In sum, these operational challenges indicate that dedicated operator training, beyond simply reading the IFU, is warranted for performance of this test to highlight potential failure modes. This recommendation for additional training is consistent with studies that have suggested that specific training in reading positive Ag RDT results may be needed to achieve high specificity [7, 8], and others that have suggested that the level of training of the operator impacts Ag RDT clinical sensitivity [17].

Our study had some limitations. We recognize that the comparator in our study was RT-PCR performed on an AN swab, as opposed to an NP swab, which is still considered the reference method by the FDA [18]. This dual AN swab study design was also used for our recent BinaxNOW study [6]. Although AN swabs have had lower sensitivity than NP swabs in some studies, the sensitivity is highly dependent on the sampling technique and assay used [19]. The dry AN swab sampling method used in this study has been shown to have similar sensitivity to paired NP swabs in transport media [13]. We also note that a recent comparison study demonstrated that Ag RDT performance with nasal mid-turbinate swabs was similar to Ag RDT performance with NP swabs [11]. The time interval between sample collection and test initiation in this study is discussed above. Finally, we recognize that our symptomatic pediatric cohort was relatively small and thus the confidence intervals on all performance estimates relatively wide.

In summary, the Access Bio CareStart Ag RDT had high sensitivity in individuals with high viral burden (Ct ≤25) and moderate sensitivity in symptomatic individuals overall. Observed specificity was lower than estimates in the manufacturer IFU, slightly lower than some other visually-read Ag RDT products on the market, and unexpectedly lower in symptomatic versus asymptomatic individuals, warranting additional study. Excellent inter-operator agreement was observed, but operational challenges indicate that operator training is warranted to highlight possible test failure modes. Centers for Disease Control and Prevention recommendations for use of Ag tests were recently updated and address use of Ag tests (with/without NAAT confirmation) in various testing scenarios based on data to date [20].

## Data Availability

All data referred to in the manuscript are available.

## Acknowledgments

We thank Kerin Milesky (Office of Emergency Preparedness, Massachusetts Department of Public Health); Samantha Phillips, Bruce Hugabone, James McGarry, Robert Martucelli, Robert Zalewski (Massachusetts Emergency Management Agency), and Christina Dispena (Massachusetts State Fire Services) for assistance with logistical and support assistance at the Lawrence General Hospital Stop the Spread site.

This publication was supported by Cooperative Agreement Number 1U60OE000103, funded by the Centers for Disease Control and Prevention through the Association of Public Health Laboratories.

The findings and conclusions in this report are those of the authors and do not necessarily represent the official position of the Centers for Disease Control and Prevention (CDC) nor the official views of the Association of Public Health Laboratories (APHL). Use of trade names and commercial sources is for identification only and does not imply endorsement by the U.S. Department of Health and Human Services or CDC.

## References

1. Johns Hopkins University. COVID-19 Dashboard by the Center for Systems Science and Engineering (CCSE) at Johns Hopkins University. https://coronavirus.jhu.edu/map.html (Accessed February 12, 2020).

2. Centers for Disease Control. CDC COVID Data Tracker. https://covid.cdc.gov/covid-data-tracker/#testing_testsperformed (Accessed February 12, 2020).

3. Food and Drug Administration. Individual EUAs for Antigen Diagnostic Tests for SARS-CoV-2. https://www.fda.gov/medical-devices/coronavirus-disease-2019-covid-19-emergency-use-authorizations-medical-devices/vitro-diagnostics-euas#individual-antigen. (Accessed February 10, 2020).

4. Krüger LJ, Gaeddert M, Köppel L, et al. Evaluation of the accuracy, ease of use and limit of detection of novel, rapid, antigen-detecting point-of-care diagnostics for SARS-CoV-2. medRxiv 2020: 2020.10.01.20203836.

5. Iglói Z, Velzing J, van Beek J, et al. Clinical evaluation of the Roche/SD Biosensor rapid antigen test with symptomatic, non-hospitalized patients in a municipal health service drive-through testing site. medRxiv 2020: 2020.11.18.20234104.

6. Pollock NR, Jacobs JR, Tran K, et al. Performance and Implementation Evaluation of the Abbott BinaxNOW Rapid Antigen Test in a High-throughput Drive-through Community Testing Site in Massachusetts. J Clin Microbiol, in press.

7. Pilarowski G, Lebel P, Sunshine S, et al. Performance characteristics of a rapid SARS-CoV-2 antigen detection assay at a public plaza testing site in San Francisco. J Infect Dis 2021.

8. Pilarowski G, Marquez C, Rubio L, et al. Field performance and public health response using the BinaxNOW TM Rapid SARS-CoV-2 antigen detection assay during community-based testing. Clin Infect Dis 2020.

9. Albert E, Torres I, Bueno F, et al. Field evaluation of a rapid antigen test (Panbio COVID-19 Ag Rapid Test Device) for COVID-19 diagnosis in primary healthcare centres. Clin Microbiol Infect 2020.

10. Prince-Guerra JL, Almendares O, Nolen LD, et al. Evaluation of Abbott BinaxNOW Rapid Antigen Test for SARS-CoV-2 Infection at Two Community-Based Testing Sites - Pima County, Arizona, November 3-17, 2020. MMWR Morb Mortal Wkly Rep 2021; 70(3): 100–5.

11. Lindner AK, Nikolai O, Kausch F, et al. Head-to-head comparison of SARS-CoV-2 antigen-detecting rapid test with self-collected anterior nasal swab versus professional-collected nasopharyngeal swab. Eur Respir Journal, in press.

12. Access Bio. Package Insert for the Access Bio CareStart COVID-19 Antigen Test. https://www.fda.gov/media/142919/download.

13. Clinical Research Sequencing Platform (CRSP), LLC at the Broad Institute of MIT and Harvard. CRSP SARS-CoV-2 Real-time Reverse Transcriptase (RT)-PCR Diagnostic Assay. https://www.fda.gov/media/139858/download.

14. Food and Drug Administration. Policy for Coronavirus Disease-2019 Tests During the Public Health Emergency (Revised), May 11, 2020. https://www.fda.gov/media/135659/download.

15. Food and Drug Administration. In Vitro Diagnostics EUAs, Antigen Template for Test Developers, October 26, 2020. https://www.fda.gov/medical-devices/coronavirus-disease-2019-covid-19-emergency-use-authorizations-medical-devices/vitro-diagnostics-euas.

16. Abbott. Package Insert for the Abbott BinaxNOW COVID-19 Ag CARD. 2020; https://www.fda.gov/media/141570/download.

17. Preliminary report from the Joint PHE Porton Down & University of Oxford SARS-CoV-2 test development and validation cell: Rapid evaluation of Lateral Flow Viral Antigen detection devices (LFDs) for mass community testing. https://www.ox.ac.uk/sites/files/oxford/media_wysiwyg/UK%20evaluation_PHE%20Porton%20Down%20%20University%20of%20Oxford_final.pdf.

18. Food and Drug Administration. FAQs on Testing for SARS-CoV-2. https://www.fda.gov/medical-devices/coronavirus-covid-19-and-medical-devices/faqs-testing-sars-cov-2.

19. Lee RA, Herigon JC, Benedetti A, Pollock NR, Denkinger C. Performance of Saliva, Oropharyngeal Swabs, and Nasal Swabs for SARS-CoV-2 Molecular Detection: A Systematic Review and Meta-analysis. J Clin Microbiol 2020, in press.

20. Centers for Disease Control. Interim Guidance for Antigen Testing for SARS-CoV-2. December 5, 2020. https://www.cdc.gov/coronavirus/2019-ncov/lab/resources/antigen-tests-guidelines.html.

