## Supplementary Materials for "Performance and Operational Evaluation of the Access Bio CareStart Rapid Antigen Test in a High-throughput Drive-through Community Testing Site in Massachusetts"

**Supplementary Methods**

Anterior Nasal Swab Collection procedure

Manufacturer-provided swabs [SteriPack Sterile Polyester Spun Swab, 3" (Lakeland, Florida)] were utilized for the entire study. The following swab collection procedure was utilized: after the participant blew their nose, the collector inserted swab #1 into the individual's right nostril and rotated the swab 5 times in a circular motion around the inside wall of the nostril for a duration of 10-15 seconds, followed by a repeat of the process in the left nostril. The swab was placed into a sterile plastic tube. Immediately thereafter, swab #2 was inserted into the individual's left nostril (always opposite from the initial swab) and the process repeated (left nostril followed by right nostril). To minimize sampling bias, the study site personnel alternated which test-specific nasal swab sample was collected first (CareStart or RT-PCR).

Testing Site

Testing was performed in a temperature-controlled mobile trailer (70.5-74.3°F) to maintain the minimal temperature (15°C) for testing per manufacturer's instructions for use (IFU). Test kits were also stored in the trailer. The testing site contained three 6' tables. Two tables were designated as "dirty" for testing purposes and one "clean" for data entry. Absorbent underpads with plastic backing (Attends ASB-2336, Greenville, NC) were placed on testing tables along with hand sanitizer, gloves, testing kits, pens, sticky notes, a continuous temperature and humidity data logger, a digital clock, and a bin for specimens to be tested. The clean table included hand sanitizer, gloves, pens, a time stamp machine, and a laptop computer for data entry. Underpads were removed daily, surfaces were disinfected with germicidal disposable wipes (PDI, Woodcliff Lake, NJ), and new underpads placed. Operators performing testing were wearing disposable gowns, N95 respirators, face shields and gloves.

Quality Control

Lot numbers were recorded for CareStart kits (boxes of 20 tests and extraction vials, and positive controls). Each study subject tested was recorded for every kit of CareStart tests used. Positive and negative controls were run on a new box at the start of each day. If an opened kit was not used in full over the course of one day, a negative control was run for the kit the following day when testing resumed.

CareStart Testing

At the collection site, anterior nasal swab specimens were collected using the manufacturer-provided swab (above) into a labeled sterile 15ml or 25ml conical tube and placed into a specimen collection bag with a CareStart intake form (which was labeled and time/date stamped at the time of collection) and extra patient labels. Specimens were then shuttled to the testing trailer by a designated individual (“runner”) by car in a closed specimen transport container. Tests were initiated as soon as possible and within an hour of specimen collection time. For each specimen, testing staff extracted the swab in the extraction buffer as per the IFU and affixed a patient label to both the outside of the extraction vial and the CareStart cartridge. The time the extracted material was applied to the test cartridge was recorded on the CareStart intake form as the test start time. The test start time and time the test was to be read (10 minutes after adding the sample) were also recorded on a sticky note and adhered to the outside of the CareStart cartridge. At the time of result reading, the tester silently recorded their result, initials, and time (between 10 and 15 minutes after adding the sample) on the CareStart intake form as the official result.

The tester then passed the cartridge to a second reader, who read the test result aloud (so that the tester could record results on the back of the intake form) and took a photo of any positive result. There was no attempt to resolve any discordance between the two readers; the first read of each test was the official result used for database and clinical reporting. If the result was determined to be positive by either reader, the reader categorized the positive line as “faint”, “medium”, or “strong” depending on the strength of the positive sample line compared to the control line of each card. Completed CareStart cartridges were discarded in biohazard waste.

RT-PCR assay details

Dry swabs (SteriPack Sterile Polyester Spun Swab, 3” (Lakeland, Florida)) transported in a sterile tube (Becton Dickinson, # BD 366408) at room temperature were resuspended in 1000 µL of Swab Preservation Buffer (Norgen Biotek, Inc) and allowed to elute for 15 minutes. Extraction and RT-PCR methods followed the EUA protocol for the CRSP SARS-CoV-2 Real-time Reverse Transcriptase (RT)-PCR Diagnostic Assay (V2) (https://www.fda.gov/media/139858/download); the target is the N2 gene with a cycle threshold cut off value of 40. A standard curve was prepared with FDA reference panel material [heat inactivated virus from BEI Resources (www.beiresources.org)] using serial dilutions (in triplicate) made in negative clinical matrix then spiked onto dry swabs, allowed to dry, and subsequently taken all the way through the process.

### Supplementary Table 1

### Access Bio CareStart results, RT-PCR Ct values, and symptoms in RT-PCR-positive adults

| Ct (N2) | Ct (RP) | CareStart Result | COVID-19 Symptoms? | Symptom Onset Date | Sore Throat | Cough | Chills | Body Aches | Shortness of Breath | Fever | Runny Nose | Congestion | Nausea | Vomiting | Diarrhea | Loss of Taste or Smell | # Days Since Symptom Onset |
|---|---|---|---|---|---|---|---|---|---|---|---|---|---|---|---|---|---|
| 24.95 | 24.57 | NEG | Asx | | | | | | | | | | | | | | n/a |
| 33.36 | 22.85 | NEG | Asx | | | | | | | | | | | | | | n/a |
| 31.76 | 21.01 | NEG | Asx | | | | | | | | | | | | | | n/a |
| 27.93 | 22.81 | NEG | Asx | | | | | | | | | | | | | | n/a |
| 33.89 | 21.36 | NEG | Asx | | | | | | | | | | | | | | n/a |
| 34.61 | 23.25 | NEG | Asx | | | | | | | | | | | | | | n/a |
| 28.67 | 25.59 | NEG | Asx | | | | | | | | | | | | | | n/a |
| 34.3 | 22.37 | NEG | Asx | | | | | | | | | | | | | | n/a |
| 35.81 | 24.12 | NEG | Asx | | | | | | | | | | | | | | n/a |
| 35.76 | 24.69 | NEG | Asx | | | | | | | | | | | | | | n/a |
| 37.23 | 23.2 | NEG | Asx | | | | | | | | | | | | | | n/a |
| 26.63 | 21.59 | NEG | Asx | | | | | | | | | | | | | | n/a |
| 30.92 | 21.51 | NEG | Asx | | | | | | | | | | | | | | n/a |
| 30.35 | 22.58 | NEG | Asx | | | | | | | | | | | | | | n/a |
| 33.26 | 22.13 | NEG | Asx | | | | | | | | | | | | | | n/a |
| 30.65 | 23.78 | NEG | Asx | | | | | | | | | | | | | | n/a |
| 36.63 | 22.78 | NEG | Asx | | | | | | | | | | | | | | n/a |
| 37.73 | 20.6 | NEG | Asx | | | | | | | | | | | | | | n/a |
| 39.05 | 17.81 | NEG | Asx | | | | | | | | | | | | | | n/a |
| 36.32 | 25.37 | NEG | Asx | | | | | | | | | | | | | | n/a |
| 34.32 | 23.89 | NEG | Asx | | | | | | | | | | | | | | n/a |
| 35.64 | 21.85 | NEG | Asx | | | | | | | | | | | | | | n/a |
| 29.7 | 24.44 | NEG | Asx | | | | | | | | | | | | | | n/a |
| 34.77 | 24.55 | NEG | Asx | | | | | | | | | | | | | | n/a |
| 29.92 | 23.35 | NEG | Asx | | | | | | | | | | | | | | n/a |
| 30.99 | 26.28 | NEG | Asx | | | | | | | | | | | | | | n/a |
| 27.38 | 24.53 | NEG | Asx | | | | | | | | | | | | | | n/a |
| 34.59 | 26.75 | NEG | Asx | | | | | | | | | | | | | | n/a |
| 27.7 | 22.64 | NEG | Asx | | | | | | | | | | | | | | n/a |
| 32.99 | 26.83 | NEG | Asx | | | | | | | | | | | | | | n/a |
| 26.67 | 21.52 | NEG | Asx | | | | | | | | | | | | | | n/a |
| 28.22 | 22.2 | NEG | Asx | | | | | | | | | | | | | | n/a |
| 28.48 | 19.64 | NEG | Asx | | | | | | | | | | | | | | n/a |
| 28.51 | 25.35 | NEG | Asx | | | | | | | | | | | | | | n/a |
| 30.55 | 22.75 | NEG | Asx | | | | | | | | | | | | | | n/a |
| 36.41 | 22.85 | NEG | Asx | | | | | | | | | | | | | | n/a |
| 32.47 | 23.63 | NEG | Asx | | | | | | | | | | | | | | n/a |
| 34.69 | 23.86 | NEG | Asx | | | | | | | | | | | | | | n/a |
| 32.62 | 21.26 | NEG | Asx | | | | | | | | | | | | | | n/a |
| 33.59 | 22.97 | NEG | Asx | | | | | | | | | | | | | | n/a |
| 34.32 | 23.75 | NEG | Asx | | | | | | | | | | | | | | n/a |
| 35.24 | 21.13 | NEG | Asx | | | | | | | | | | | | | | n/a |
| 35.55 | 23.88 | NEG | Asx | | | | | | | | | | | | | | n/a |
| 30.41 | 23.8 | NEG | Asx | | | | | | | | | | | | | | n/a |
| 26.65 | 21.23 | NEG | Asx | | | | | | | | | | | | | | n/a |
| 34.47 | 22.41 | NEG | Asx | | | | | | | | | | | | | | n/a |
| 34.07 | 22.28 | NEG | Asx | | | | | | | | | | | | | | n/a |
| 34.4 | 27.22 | NEG | Asx | | | | | | | | | | | | | | n/a |
| 30.29 | 23.7 | NEG | Asx | | | | | | | | | | | | | | n/a |
| 27.91 | 23.4 | NEG | Asx | | | | | | | | | | | | | | n/a |
| 26.94 | 22.33 | NEG | Asx | | | | | | | | | | | | | | n/a |
| 26.93 | 20.95 | NEG | Asx | | | | | | | | | | | | | | n/a |
| 27.79 | 25 | NEG | Asx | | | | | | | | | | | | | | n/a |
| 29.76 | 20.61 | NEG | Asx | | | | | | | | | | | | | | n/a |
| 32.46 | 24.46 | NEG | Asx | | | | | | | | | | | | | | n/a |
| 27.35 | 21.59 | NEG | Asx | | | | | | | | | | | | | | n/a |
| 29.62 | 24.25 | NEG | Asx | | | | | | | | | | | | | | n/a |
| 23.18 | 23.38 | NEG | Asx | | | | | | | | | | | | | | n/a |
| 21.91 | 22.68 | NEG | Asx | | | | | | | | | | | | | | n/a |
| 34.68 | 23.38 | NEG | Asx | | | | | | | | | | | | | | n/a |
| 28.26 | 24.32 | NEG | Asx | | | | | | | | | | | | | | n/a |
| 29.9 | 24.29 | NEG | Asx | | | | | | | | | | | | | | n/a |
| 27.91 | 25.82 | NEG | Asx | | | | | | | | | | | | | | n/a |
| 31.44 | 23.77 | NEG | Asx | | | | | | | | | | | | | | n/a |
| 20.1 | 20.4 | POS | Asx | | | | | | | | | | | | | | n/a |
| 20.79 | 22.44 | POS | Asx | | | | | | | | | | | | | | n/a |
| 18.51 | 22.46 | POS | Asx | | | | | | | | | | | | | | n/a |
| 31.35 | 26.2 | POS | Asx | | | | | | | | | | | | | | n/a |
| 19.15 | 23.86 | POS | Asx | | | | | | | | | | | | | | n/a |
| 17.34 | 24.02 | POS | Asx | | | | | | | | | | | | | | n/a |
| 17.08 | 23.56 | POS | Asx | | | | | | | | | | | | | | n/a |
| 13.96 | 22.88 | POS | Asx | | | | | | | | | | | | | | n/a |
| 20.45 | 21.9 | POS | Asx | | | | | | | | | | | | | | n/a |
| 15.13 | 25.65 | POS | Asx | | | | | | | | | | | | | | n/a |
| 30.15 | 22.31 | POS | Asx | | | | | | | | | | | | | | n/a |
| 17.4 | 23.92 | POS | Asx | | | | | | | | | | | | | | n/a |
| 26.2 | 27.71 | POS | Asx | | | | | | | | | | | | | | n/a |
| 20.09 | 24.63 | POS | Asx | | | | | | | | | | | | | | n/a |
| 17.47 | 24.13 | POS | Asx | | | | | | | | | | | | | | n/a |
| 24.81 | 23.71 | POS | Asx | | | | | | | | | | | | | | n/a |
| 17.92 | 23.84 | POS | Asx | | | | | | | | | | | | | | n/a |
| 22.76 | 23.29 | POS | Asx | | | | | | | | | | | | | | n/a |
| 25.47 | 28 | POS | Asx | | | | | | | | | | | | | | n/a |
| 17.89 | 21.26 | POS | Asx | | | | | | | | | | | | | | n/a |
| 29.49 | 23.34 | POS | Asx | | | | | | | | | | | | | | n/a |
| 15.54 | 19.51 | POS | Asx | | | | | | | | | | | | | | n/a |
| 28.99 | 24.59 | POS | Asx | | | | | | | | | | | | | | n/a |
| 28.41 | 24.24 | POS | Asx | | | | | | | | | | | | | | n/a |
| 15.67 | 21.97 | POS | Asx | | | | | | | | | | | | | | n/a |
| 15.31 | 23.37 | POS | Asx | | | | | | | | | | | | | | n/a |
| 23.61 | 19.32 | POS | Asx | | | | | | | | | | | | | | n/a |
| 26.5 | 22.63 | POS | Asx | | | | | | | | | | | | | | n/a |
| 22.62 | 25.21 | POS | Asx | | | | | | | | | | | | | | n/a |

| | | | | | | | | | | | | | | | | | |
|---|---|---|---|---|---|---|---|---|---|---|---|---|---|---|---|---|---|
| 24.41 | - | POS | Asx | | | | | | | | | | | | | | n/a |
| 12.47 | 21.43 | POS | Asx | | | | | | | | | | | | | | n/a |
| 23.18 | 24.56 | POS | Asx | | | | | | | | | | | | | | n/a |
| 24.41 | 25.64 | POS | Asx | | | | | | | | | | | | | | n/a |
| 14.44 | 21.15 | POS | Asx | | | | | | | | | | | | | | n/a |
| 21.5 | 24.19 | POS | Asx | | | | | | | | | | | | | | n/a |
| 22.83 | 23.52 | POS | Asx | | | | | | | | | | | | | | n/a |
| 21.58 | 26.71 | POS | Asx | | | | | | | | | | | | | | n/a |
| 27.19 | 24.61 | POS | Asx | | | | | | | | | | | | | | n/a |
| 21.91 | 21.77 | POS | Asx | | | | | | | | | | | | | | n/a |
| 19.98 | 25.2 | POS | Asx | | | | | | | | | | | | | | n/a |
| 17.22 | 22.78 | POS | Asx | | | | | | | | | | | | | | n/a |
| 19.89 | 23.87 | POS | Asx | | | | | | | | | | | | | | n/a |
| 25.33 | 22.72 | POS | Asx | | | | | | | | | | | | | | n/a |
| 19.87 | 20.88 | POS | Asx | | | | | | | | | | | | | | n/a |
| 29.28 | 24.49 | POS | Asx | | | | | | | | | | | | | | n/a |
| 22.84 | 26.33 | POS | Asx | | | | | | | | | | | | | | n/a |
| 24.44 | 24.72 | POS | Asx | | | | | | | | | | | | | | n/a |
| 15.21 | 21.72 | POS | Asx | | | | | | | | | | | | | | n/a |
| 18.05 | 23.7 | POS | Asx | | | | | | | | | | | | | | n/a |
| 21.53 | 24.85 | POS | Asx | | | | | | | | | | | | | | n/a |
| 18.74 | 22.46 | POS | Asx | | | | | | | | | | | | | | n/a |
| 17.39 | 22.39 | POS | Asx | | | | | | | | | | | | | | n/a |
| 15.05 | 23.17 | POS | Asx | | | | | | | | | | | | | | n/a |
| 27.77 | 23.31 | POS | Asx | | | | | | | | | | | | | | n/a |
| 18.41 | 24.96 | POS | Asx | | | | | | | | | | | | | | n/a |
| 30.53 | 21.95 | POS | Asx | | | | | | | | | | | | | | n/a |
| 16.52 | 25.91 | POS | Asx | | | | | | | | | | | | | | n/a |
| 31.79 | 27.22 | POS | Asx | | | | | | | | | | | | | | n/a |
| 17.19 | 26.31 | POS | Asx | | | | | | | | | | | | | | n/a |
| 19.87 | 20.59 | POS | Asx | | | | | | | | | | | | | | n/a |
| 14.11 | 23.89 | POS | Asx | | | | | | | | | | | | | | n/a |
| 21.23 | 26.53 | POS | Asx | | | | | | | | | | | | | | n/a |
| 16.26 | 22.54 | POS | Asx | | | | | | | | | | | | | | n/a |
| 22.82 | 22.93 | POS | Asx | | | | | | | | | | | | | | n/a |
| 24.32 | 24.6 | NEG | Sx | 1/13/21 | | | | | | | Y | Y | | | | | 0 |
| 27.57 | 26.5 | NEG | Sx | 1/10/21 | Y | | | Y | | | | | | | | | 1 |
| 34.49 | 22.01 | NEG | Sx | 1/12/21 | Y | Y | Y | | | | | | | | | | 1 |
| 31.47 | 22.84 | NEG | Sx | 1/12/21 | | | | Y | | | | | | | | | 2 |
| 32.39 | 21.68 | NEG | Sx | 1/10/21 | Y | | | | | | | | | | | | 3 |
| 32.34 | 21.41 | NEG | Sx | 1/9/21 | Y | | | | | | | Y | | | | | 4 |
| 30.94 | 28.45 | NEG | Sx | 1/18/21 | | | Y | Y | | | | Y | | | | | 4 |
| 34.66 | 19.56 | NEG | Sx | 1/7/21 | | Y | Y | Y | | | Y | | Y | | | | 6 |
| 18.46 | 25.78 | POS | Sx | 1/14/21 | Y | Y | | Y | | | Y | Y | | | | | 0 |
| 18.99 | 27.38 | POS | Sx | 1/22/21 | | | Y | Y | | Y | Y | | | | | | 0 |
| 16.5 | 24.87 | POS | Sx | 1/13/21 | | Y | | | | | | | | | | | 0 |
| 20.54 | 26.15 | POS | Sx | 1/20/21 | Y | | | | | Y | | | | | | | 0 |
| 20.52 | 25.3 | POS | Sx | 1/12/21 | Y | Y | | | | | Y | Y | | | | | 1 |
| 20.35 | 22.11 | POS | Sx | 1/12/21 | Y | | Y | | | | | Y | | | | Y | 1 |
| 14.22 | 25.06 | POS | Sx | 1/20/21 | Y | | | Y | | | | | | | | | 1 |
| 20.42 | 23.12 | POS | Sx | 1/13/21 | | | | | | Y | Y | | | | | | 1 |
| 18.02 | 25.18 | POS | Sx | 1/18/21 | | | Y | Y | | | | | | | | | 1 |
| 11.15 | 19.98 | POS | Sx | 1/19/21 | | | | | | | Y | | | | | | 1 |
| 24.67 | 23.37 | POS | Sx | 1/13/21 | | | | | | | Y | Y | | | | | 1 |
| 19.76 | 20.23 | POS | Sx | 1/18/21 | | Y | | | | | | | | | | | 2 |
| 25.25 | 25.26 | POS | Sx | 1/17/21 | Y | Y | Y | Y | | Y | | | | | | | 2 |
| 17.57 | 20.68 | POS | Sx | 1/18/21 | | Y | | Y | | Y | Y | Y | | | | Y | 2 |
| 28.57 | 25.71 | POS | Sx | 1/12/21 | Y | Y | Y | Y | Y | Y | Y | Y | | | | Y | 2 |
| 26.23 | 26.53 | POS | Sx | 1/9/21 | Y | Y | | Y | Y | Y | Y | Y | | | | Y | 2 |
| 13.27 | 20.3 | POS | Sx | 1/9/21 | Y | Y | Y | | | | Y | Y | | | | | 2 |
| 20.66 | 30.69 | POS | Sx | 1/9/21 | | Y | | | | | | Y | | | | | 2 |
| 17.11 | 27.31 | POS | Sx | 1/19/21 | Y | | Y | Y | | | Y | | | | | | 2 |
| 20.78 | 19.35 | POS | Sx | 1/16/21 | | Y | Y | Y | | | | | | | | | 3 |
| 24.49 | 22.44 | POS | Sx | 1/18/21 | | | | | | | Y | | | | | | 3 |
| 17.94 | 24.84 | POS | Sx | 1/8/21 | Y | | | | | | Y | Y | | | Y | | 3 |
| 22.8 | 25.21 | POS | Sx | 1/16/21 | | | | | | | | | | | | Y | 3 |
| 19.13 | 22.43 | POS | Sx | 1/18/21 | | | | | | | | Y | | | | | 3 |
| 23.14 | 26.72 | POS | Sx | 1/17/21 | Y | Y | Y | Y | Y | Y | | Y | Y | Y | Y | | 3 |
| 18.36 | 24.44 | POS | Sx | 1/17/21 | | Y | | Y | | Y | | | | | | | 3 |
| 13.5 | 22.67 | POS | Sx | 1/16/21 | | Y | | | | | Y | | | | | Y | 3 |
| 23.61 | 22.95 | POS | Sx | 1/16/21 | | Y | Y | Y | Y | Y | Y | | | | | | 3 |
| 12.7 | 21.82 | POS | Sx | 1/10/21 | Y | Y | Y | | | | | | | | | | 3 |
| 15.21 | 24.83 | POS | Sx | 1/17/21 | | Y | | Y | Y | Y | | | | | | | 3 |
| 17.48 | 23.87 | POS | Sx | 1/7/21 | Y | Y | Y | Y | | Y | Y | Y | | | Y | Y | 4 |
| 22.3 | 25.51 | POS | Sx | 1/7/21 | Y | Y | | Y | | | Y | Y | Y | | Y | | 4 |
| 15.34 | 21.63 | POS | Sx | 1/9/21 | Y | | | Y | | Y | | | | | Y | | 4 |
| 16.41 | 23.83 | POS | Sx | 1/8/21 | | Y | | Y | | | Y | Y | | | | | 4 |
| 23.41 | 25.7 | POS | Sx | 1/8/21 | | | Y | Y | | | Y | Y | | | | Y | 4 |
| 18.49 | 26.12 | POS | Sx | 1/8/21 | Y | Y | | Y | | | Y | | | | | | 4 |
| 19.79 | 23.45 | POS | Sx | 1/14/21 | | Y | | | | Y | | | | | | | 5 |
| 14.55 | 22.67 | POS | Sx | 1/6/21 | | Y | | Y | | Y | Y | | | | | | 5 |
| 20.05 | 22.85 | POS | Sx | 1/16/21 | Y | Y | | Y | Y | | | | | | | | 5 |
| 19.36 | 20.62 | POS | Sx | 1/14/21 | Y | Y | Y | Y | Y | N | Y | Y | N | N | N | N | 6 |
| 22.26 | 26.3 | POS | Sx | 1/14/21 | | | | Y | Y | | | Y | | | | | 6 |
| 18.66 | 23.4 | POS | Sx | 1/13/21 | | Y | | Y | Y | | Y | | Y | Y | Y | | 6 |
| 25.14 | 26.27 | POS | Sx | 1/13/21 | | Y | | Y | | | | Y | | | | | 7 |
| 31.22 | 25.09 | NEG | Sx7+ | 1/4/21 | Y | | | | | | Y | | | | | | 10 |
| 35.02 | 25.01 | NEG | Sx7+ | 1/4/21 | | | | | Y | | Y | | | | | | 10 |
| 32.6 | 23.7 | NEG | Sx7+ | 1/4/21 | Y | Y | | | | | Y | | | | | | 10 |
| 31.17 | 23.9 | NEG | Sx7+ | 1/7/21 | | Y | | | | | | | | | | | 14 |
| 26.93 | 28 | NEG | Sx7+ | 1/4/21 | Y | Y | | Y | | | Y | Y | Y | | | | 15 |
| 33.65 | 26.48 | NEG | Sx7+ | 1/5/21 | | | | | | | | Y | | | | | 16 |
| 34.75 | 26.46 | NEG | Sx7+ | 12/27/20 | Y | Y | Y | Y | | | Y | Y | | | | Y | 23 |
| 20.57 | 23.51 | POS | Sx7+ | 1/1/21 | | | | | | | | | | | | Y | 11 |
| 14.43 | 19.66 | POS | Sx7+ | 1/6/21 | | Y | Y | Y | Y | | Y | | | | | | 14 |

RT-PCR, real-time reverse transcription polymerase chain reaction; F, female; M, male; POS, positive; NEG, negative; Ct, cycle threshold; N2, SARS-CoV-2 target; RP, host control target; ASx, asymptomatic; Sx, symptomatic; n/a, not applicable

**Supplementary Table 2**

**Access Bio CareStart results, RT-PCR Ct values, and symptoms in RT-PCR-positive children**

| Ct (N2) | Ct (RP) | CareStart Result | COVID-19 Symptoms? | Symptom Onset Date | Sore Throat | Cough | Chills | Body Aches | Shortness of Breath | Fever | Runny Nose | Congestion | Nausea | Vomiting | Diarrhea | Loss of Taste or Smell | # Days Since Symptom Onset |
|---|---|---|---|---|---|---|---|---|---|---|---|---|---|---|---|---|---|
| 29.96 | 26.16 | NEG | ASx | | | | | | | | | | | | | | n/a |
| 30.77 | 26.34 | NEG | ASx | | | | | | | | | | | | | | n/a |
| 34.39 | 20.8 | NEG | ASx | | | | | | | | | | | | | | n/a |
| 32.05 | 22.43 | NEG | ASx | | | | | | | | | | | | | | n/a |
| 28.09 | 23.37 | NEG | ASx | | | | | | | | | | | | | | n/a |
| 34.43 | 22.09 | NEG | ASx | | | | | | | | | | | | | | n/a |
| 28.75 | 21.62 | NEG | ASx | | | | | | | | | | | | | | n/a |
| 31.65 | 22.16 | NEG | ASx | | | | | | | | | | | | | | n/a |
| 32.25 | 21.06 | NEG | ASx | | | | | | | | | | | | | | n/a |
| 36.73 | 20.38 | NEG | ASx | | | | | | | | | | | | | | n/a |
| 34.95 | 23.23 | NEG | ASx | | | | | | | | | | | | | | n/a |
| 35.48 | 23.37 | NEG | ASx | | | | | | | | | | | | | | n/a |
| 28.81 | 24.7 | NEG | ASx | | | | | | | | | | | | | | n/a |
| 27.74 | 21.65 | NEG | ASx | | | | | | | | | | | | | | n/a |
| 32.9 | 22.36 | NEG | ASx | | | | | | | | | | | | | | n/a |
| 32 | 27.23 | NEG | ASx | | | | | | | | | | | | | | n/a |
| 34.29 | 22.37 | NEG | ASx | | | | | | | | | | | | | | n/a |
| 34.64 | 22.36 | NEG | ASx | | | | | | | | | | | | | | n/a |
| 27.85 | 23.64 | POS | ASx | | | | | | | | | | | | | | n/a |
| 22.61 | 26.97 | POS | ASx | | | | | | | | | | | | | | n/a |
| 26.08 | 21.33 | POS | ASx | | | | | | | | | | | | | | n/a |
| 19.9 | 21.12 | POS | ASx | | | | | | | | | | | | | | n/a |
| 31.81 | 23.45 | POS | ASx | | | | | | | | | | | | | | n/a |
| 26.3 | 22 | POS | ASx | | | | | | | | | | | | | | n/a |
| 24.16 | 24.74 | POS | ASx | | | | | | | | | | | | | | n/a |
| 31.74 | 20.19 | POS | ASx | | | | | | | | | | | | | | n/a |
| 18.51 | 24.67 | POS | ASx | | | | | | | | | | | | | | n/a |
| 16.01 | 23.3 | POS | ASx | | | | | | | | | | | | | | n/a |
| 22.77 | 22.61 | POS | ASx | | | | | | | | | | | | | | n/a |
| 15.44 | 20.14 | POS | ASx | | | | | | | | | | | | | | n/a |
| 13.7 | 23.09 | POS | ASx | | | | | | | | | | | | | | n/a |
| 15.12 | 23.28 | POS | ASx | | | | | | | | | | | | | | n/a |
| 18.04 | 23.59 | POS | ASx | | | | | | | | | | | | | | n/a |
| 18.59 | 22.03 | POS | ASx | | | | | | | | | | | | | | n/a |
| 23.88 | 27.23 | POS | ASx | | | | | | | | | | | | | | n/a |
| 19.95 | 23.95 | POS | ASx | | | | | | | | | | | | | | n/a |
| 16.81 | 25.38 | POS | ASx | | | | | | | | | | | | | | n/a |
| 30.83 | 24.85 | NEG | Sx | 1/12/21 | Y | Y | | | Y | | Y | | | | | | 1 |
| 22.57 | 25.33 | POS | Sx | 1/9/21 | Y | | | | | | Y | | | | | | 2 |
| 26.59 | 27.14 | POS | Sx | 1/19/21 | | Y | | Y | | | | Y | Y | Y | | | 3 |
| 22.68 | 21.28 | POS | Sx | 1/12/21 | | | | | | | | | | Y | | | 2 |
| 19.25 | 24.04 | POS | Sx | 1/16/21 | | Y | | Y | | Y | | | | | | Y | 3 |
| 14.76 | 24.93 | POS | Sx | 1/17/21 | Y | Y | | Y | | Y | Y | | | | | | 2 |
| 21.88 | 24.97 | POS | Sx | 1/16/21 | | | | Y | | | | Y | | | | Y | 3 |
| 34.52 | 23.61 | NEG | Sx7+ | 1/3/21 | | Y | | | | Y | Y | Y | Y | | | | 10 |
| 20.28 | 25.88 | POS | Sx7+ | 1/1/21 | Y | Y | | | Y | Y | Y | Y | | | | | 10 |

RT-PCR, real-time reverse transcription polymerase chain reaction; F, female; M, male; POS, positive; NEG, negative; Ct, cycle threshold; N2, SARS-CoV-2 target; RP, host control target; ASx, asymptomatic; Sx, symptomatic; n/a, not applicable

**Supplementary Table 3**

**Distribution of positive and negative CareStart and RT-PCR results across clinical subgroups**

| Asymptomatic (Children: ≤18 y) | | | |
|---|---|---|---|
| | RT-PCR | | |
| CareStart Ag | Positive | Negative | Total |
| Positive | 19 | 4 | 23 |
| Negative | 18 | 180 | 198 |
| Total (16.7% prevalence) | 37 | 184 | 221 |

| Asymptomatic (Adults: >18 y) | | | |
|---|---|---|---|
| | RT-PCR | | |
| CareStart Ag | Positive | Negative | Total |
| Positive | 64 | 8 | 72 |
| Negative | 64 | 900 | 964 |
| Total (12.4% prevalence) | 128 | 908 | 1036 |

| Symptomatic ≤5 days (Children: ≤18 y) | | | |
|---|---|---|---|
| | RT-PCR | | |
| CareStart Ag | Positive | Negative | Total |
| Positive | 6 | 2 | 8 |
| Negative | 1 | 17 | 18 |
| Total (26.9% prevalence) | 7 | 19 | 26 |

| Symptomatic ≤5 days (Adults: >18 y) | | | |
|---|---|---|---|
| | RT-PCR | | |
| CareStart Ag | Positive | Negative | Total |
| Positive | 39 | 3 | 42 |
| Negative | 7 | 103 | 110 |
| Total (30.3% prevalence) | 46 | 106 | 152 |

| Symptomatic ≤7 days (Children: ≤18 y) | | | |
|---|---|---|---|
| | RT-PCR | | |
| CareStart Ag | Positive | Negative | Total |
| Positive | 6 | 3 | 9 |
| Negative | 1 | 17 | 18 |
| Total (25.9% prevalence) | 7 | 20 | 27 |

| Symptomatic ≤7 days (Adults: >18 y) | | | |
|---|---|---|---|
| | RT-PCR | | |
| CareStart Ag | Positive | Negative | Total |
| Positive | 43 | 3 | 46 |
| Negative | 8 | 115 | 123 |
| Total (30.2% prevalence) | 51 | 118 | 169 |

| Symptomatic >7 days (Children ≤18 y) | | | |
|---|---|---|---|
| | RT-PCR | | |
| CareStart Ag | Positive | Negative | Total |
| Positive | 1 | 0 | 1 |
| Negative | 1 | 3 | 4 |
| Total (40.0% prevalence) | 2 | 3 | 5 |

| Symptomatic >7 days (Adults >18 y) | | | |
|---|---|---|---|
| | RT-PCR | | |
| CareStart Ag | Positive | Negative | Total |
| Positive | 2 | 3 | 5 |
| Negative | 7 | 28 | 35 |
| Total (22.5% prevalence) | 9 | 31 | 40 |

RT-PCR, real-time reverse transcription polymerase chain reaction; Y, years; Ag, Antigen

**Supplementary Table 4**

**Performance of the Access Bio CareStart versus RT-PCR with varied Ct value cutoffs**

| | All Ct Values | | | | Ct values ≤35 | | | | Ct values ≤30 | | | | Ct values ≤25 | | | |
|---|---|---|---|---|---|---|---|---|---|---|---|---|---|---|---|---|
| | N | TP (+/+) | FN (+/-) | Sensitivity | N | TP (+/+) | FN (+/-) | Sensitivity | N | TP (+/+) | FN (+/-) | Sensitivity | N | TP (+/+) | FN (+/-) | Sensitivity |
| Combined All | 234 | 135 | 99 | 57.7% | 220 | 135 | 85 | 61.4% | 162 | 129 | 33 | 79.6% | 107 | 103 | 4 | 96.3% |
| All Children (All Sx, ASx) | 46 | 26 | 20 | 56.5% | 44 | 26 | 18 | 59.1% | 29 | 24 | 5 | 82.8% | 12 | 12 | 0 | 100.0% |
| All Adult (All Sx, ASx) | 188 | 109 | 79 | 58.0% | 176 | 109 | 67 | 61.9% | 133 | 105 | 28 | 78.9% | 95 | 91 | 4 | 95.8% |
| Sx (≤5D) Children | 7 | 6 | 1 | 85.7% | 7 | 6 | 1 | 85.7% | 6 | 6 | 0 | 100.0% | 5 | 5 | 0 | 100.0% |
| Sx (≤5D) Adult | 46 | 39 | 7 | 84.8% | 46 | 39 | 7 | 84.8% | 41 | 39 | 2 | 92.9% | 37 | 36 | 1 | 97.3% |
| Sx (≤7D) Children | 7 | 6 | 1 | 85.7% | 7 | 6 | 1 | 85.7% | 6 | 6 | 0 | 100.0% | 5 | 5 | 0 | 100.0% |
| Sx (≤7D) Adult | 51 | 43 | 8 | 84.3% | 51 | 43 | 8 | 84.3% | 45 | 43 | 2 | 95.6% | 40 | 39 | 1 | 97.5% |
| ASx Children | 37 | 19 | 18 | 51.4% | 34 | 19 | 15 | 55.9% | 22 | 17 | 5 | 77.3% | 14 | 14 | 0 | 100.0% |
| ASx Adult | 128 | 64 | 64 | 50.0% | 117 | 64 | 53 | 54.7% | 85 | 60 | 25 | 70.6% | 53 | 50 | 3 | 94.3% |

RT-PCR, real-time reverse transcription polymerase chain reaction; Ct, cycle threshold; TP, true positive (RT-PCR positive/CareStart positive); FN, false negative (RT-PCR positive/CareStart negative); Sens, sensitivity; Sx, symptomatic; ASx, asymptomatic; D, days
